# The impact of leadership on the nursing workforce during the COVID-10 pandemic

**DOI:** 10.1101/2021.05.18.21256802

**Authors:** Natasha Phillips, Luke Hughes, Cecilia Vindrola-Padros, Anika Petrella, Lorna A Fern, Flo Panel-Coates, Rachel M Taylor

## Abstract

**Aims:** To determine how the learning about protective factors from previous pandemics were implemented and the impact of this on nurses’ experience.

**Background:** The COVID-19 pandemic led to systemic change within healthcare settings and demands placed on frontline nurses has been overwhelming. Lessons learned from previous pandemics indicate that clear communication and strong visible leadership can mitigate the impact stressful events may have on nurses. Conversely, a lack of clear leadership and regulatory protocols in times of crisis can lead to an increase in psychological distress for nurses.

**Design:** Secondary analysis of semi-structured interview transcripts.

**Methods:** Secondary data analysis was conducted on data collected during a hospital-wide evaluation of barriers and facilitators to changes implemented to support the surge of COVID-19 related admissions in wave one of the pandemic. Participants represented three-levels of leadership: whole trust (n=17), division (n=7), ward/department-level (n=8), and individual nurses (n=16). Data were collected through semi-structured video interviews between May and July 2020. Interviews were analysed using Framework analysis.

**Results:** Key changes that were implemented in wave one reported at whole trust level included: a new acute staffing level, redeploying nurses, increasing the visibility of nursing leadership, new staff wellbeing initiatives, new roles created to support families and various training initiatives. Two main themes emerged from the interviews at division, ward/department and individual nurse level: impact of leadership, and impact on the delivery of nursing care.

**Conclusions:** Leadership through a crisis is essential for the protective effect of nurses’ emotional wellbeing. While nursing leadership was made more visible during wave one of the pandemic and processes were in place to increase communication, system-level challenges resulting in negative experiences existed. By identifying these challenges, it has been possible to overcome them during wave two by employing different leadership styles, to support nurse wellbeing

## Introduction

COVID-19 is an outbreak of a novel coronavirus disease which was declared a global pandemic by the World Health Organisation (WHO) in the early months of 2020. The disease has proven difficult to manage due to its high level of infection and transferability. The virus can be spread through, among other methods, person-person contact and aerosol transmissions via respiratory droplets (Peng et al., 2020). Due to its highly transmittable nature, and the impact it has on those who become infected, management of the disease has resulted in widespread restrictions globally, with particular relevance to establishing protocols in healthcare to combat the spread of the virus (Choudhury et al., 2020). Those working on the frontline of healthcare services were at increased risk of infection and were asked to work in ways that were unclear, unfamiliar or contrary to the normative avenues of their roles (Barello, Palamenghi & Graffinga, 2020, Xiao et al, 2020). As with many previous viral outbreaks, or other catastrophic scenarios such as wars, acts of terrorism, or natural disasters, nurses are a core element of the frontline response (Maunder et al., 2008, Xue et al., 2020). Past viral outbreaks, such as severe acute respiratory syndrome (SARS), Middle East respiratory syndrome (MERS) and Ebola, have highlighted the level of exposure nursing staff experience to both the disease and associated stressful stimuli, which often results in high levels of systemic burnout and chronic psychological distress (Maunder et al., 2004, McAlonan et al, 2007, Ulrich et al., 2014, Lee, Hong & Park, 2020, Vera San Juan et al., 2021). Likewise, investigations into the effects of natural disasters on nurses have highlighted many areas that have been impacted by these sudden drastic changes to their roles (Xue et al., 2020). Understanding how nursing staff are affected by changes in the delivery of care in exceptional circumstances is important to inform how the leadership of organisational change in a crisis can be managed so as to protect nurses’ emotional wellbeing whilst also ensuring the safe delivery of care (Chaudhry & Raza, 2020, Nyashanu, Pfende, & Ekpenyong, 2020, Vindrola-Padros et al.,2020, Vera San Juan et al., 2020, Aughterson et al., 2021, Hoernke et al., 2021, Regenold & Vindrola-Padros, 2021).

## Background

Times of great systemic change within a healthcare setting can have negative consequences on nurses working on the frontline (Brown, Zijlstra & Lyons, 2006). There is indeed a lengthy history of nurses feeling as though restructuring at an organisational level often fails to account for their experiences, and that changes may come with many impractical implications on their roles as a result of their voices not being heard by those within the managerial hierarchy (Aiken et al., 2001, Taylor et al., 1999). Previous studies have shown that sudden changes to the way staff are asked to work are associated with low job satisfaction, high job stress and lower quality of life for nurses who describe these transitions, and were characterised by poor communication, insufficient information provision and increased workloads with little access to necessary resources (Baumann et al., 2001).

Frustrations may manifest at a personal level in terms of nurses not feeling listened to when they voice concerns about how intended changes will affect their work, but also at a more widespread organisational level, where executive management evokes an authoritarian approach to restructures, or changes appear to have been made with limited foresight into the long-term consequences for the service (Brown, Zijlstra & Lyons, 2006). Alongside these organisational elements, perceptions of danger and threat in high alert scenarios can impact on frontline staff wellbeing, which can be directly mediated by organisational leadership. The higher the perception of a threat, the more likely it is to result in event related distress, and chronic or repeated exposure more than 6-months, is likely to result in long-term impacts on the quality of care provided by staff (Anderson, Ziedonist & Najavits, 2014, Rybojad et al., 2019).

Clear communication and strong visible leadership are elements associated with mitigating the impact high stress events can have on healthcare staff. A lack of clear leadership and regulatory protocols in times of crisis can lead to an increase in psychological distress for nurses (Li et al., 2017, Mao et al., 2018). In particular, the decisions of nurse leaders (sisters and matrons), and consequently how these decisions are communicated to their teams, have direct impact on the quality of nursing provided during a crisis, and consequently, impact on patient safety (Xue et al., 2020). However, it is important to note that nurse leaders may likewise be affected by the choices and communications of their executive superiors. Poor communication from organisational leaders can leave nurse leaders feeling frustrated and anxious providing guidance to their teams which they feel are vague or subject to frequent change in a crisis (Nasrabadi et al., 2007, Xue et al., 2020). Nursing leaders therefore have significant emotional weight added to their roles, both in managing their teams, managing the crisis itself, and engaging with the organisation at large. They are often required to adapt quickly to these uncertain circumstances to continue to ensure reliable and safe care for both their staff and the patient base as a whole (Aquilia et al., 2020). The tension of balancing these priorities in decision making has been recognised as central to the daily practice of nurse leadership (Phillips & Norman 2020). Arguably this is a pre-existing challenge of nurse leaders working in complex healthcare systems that is exacerbated in a rapidly evolving situation such as COVID-19 where there is a higher degree of ambiguity and uncertainty.

### The present study

The emotional and psychological impact of COVID-19 has already been shown to have a higher impact on nurses and other healthcare staff, more so than any other recent pandemics or natural disasters (Barello, Palamenghi & Graffinga, 2020, Chen et al., 2020, Xiao et al., Xue et al., 2020, 2020, Zhang et al., 2020; Zhou et al., 2020). It is likely that the organisational elements which have caused stress in previous crisis situations reoccur again. It is also likely that the daily stress of balancing competing needs to ensure the effective delivery of care is exacerbated in a rapidly evolving situation. A rapid review of the psychological effect of viral outbreaks on healthcare workers identified a series of protective factors, such as clear communication, leadership and access to personal protective equipment (PPE), as well as access to psychological support, provisions of food and necessities, adequate shift patterns, options of alternative accommodation and access to up-to-date training and education (Kisely et al., 2020). These strategies were advised retrospectively by the review for all healthcare settings moving forward to help to protect frontline staff. We undertook an evaluation of a large central London hospital at the beginning of the pandemic to determine the impact on nursing of the changes that were implemented to accommodate the increase in COVID-19 related admissions. Given the knowledge of how past pandemics have affected the nurse leaders and nurses working at the frontline, this was secondary analysis of data collected in the evaluation to determine how the learning about protective factors from previous pandemics were implemented and the impact of this on nurses’ experience. We also aimed to identify if there were more complex occupational challenges which needed to be further addressed to support nursing wellbeing in future waves.

## Methods

### Study design

This was secondary analysis of data collected for a service evaluation project, which used semi-structured interviews based on a vertical slicing framework, i.e., interviews were undertaken at the different layers of organisational leadership.

### Participants and setting

Data were generated from an evaluation of the changes implemented across a single National Health Service (NHS) university hospital to provide efficient and safe healthcare during the first wave of the COVID-19 pandemic. This was a large inner-city hospital comprising of facilities on thirteen sites, with 665 inpatient beds, employing over 9,700 members of staff, of whom nearly 3,500 were nurses and midwives (UCLH, 2020).

Recruitment to the evaluation was purposeful and conducted in three phases, involving four tiers of leadership (Figure 1). The initial sample were recruited in May 2020 and comprised of departmental leads, deputy chief nurses and other senior members of the management team who led the changes to the delivery of care across the whole Trust. The second wave of recruitment involved matrons, sisters/charge nurses and senior clinical nurse specialist (CNS) and senior clinical research nurses (CRN) recruited in May/June 2020. Matrons represented leadership across a division and the latter groups were nurses responsible for leading a team. The final wave were recruited in June/July 2020 and represented individual nurses. This was a convenience sample of CNS/CRNs who were either redeployed or covered work for colleagues who were redeployed. The aim was to recruit representation from operational leads for all the areas where there were changes, and ten participants for each of the other groups.

**Figure 1:**
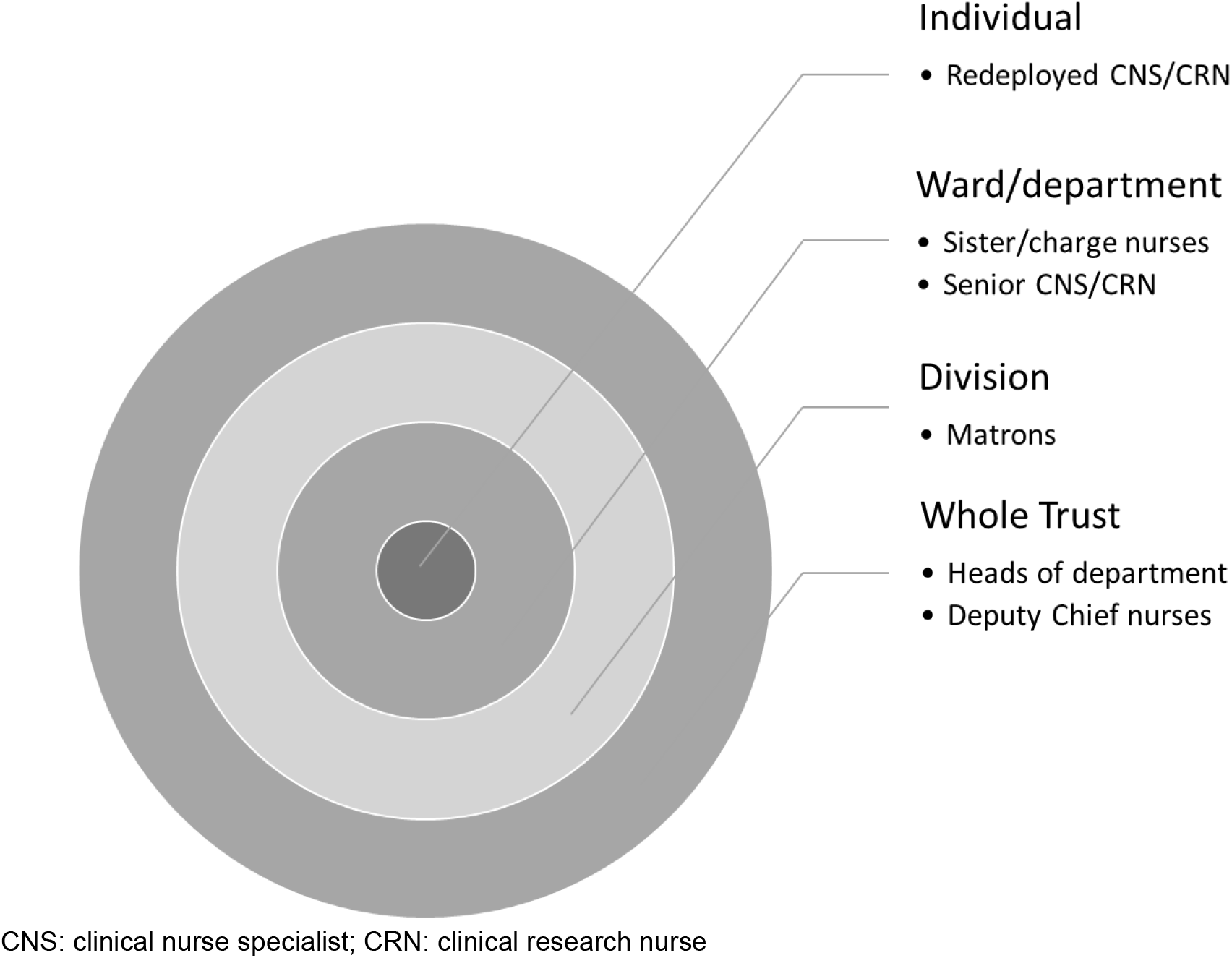
Summary of the participants in the study

Participants were invited to participate through an introductory email from the Nursing and Midwifery Leadership Team, with directions to contact the research team directly to ensure anonymity. While this was classified as service evaluation it was conducted in accordance with the UK Framework for Health and Social Care Research (Health Research Authority, 2017). The evaluation was approved by the Nursing and Midwifery Leadership Team, who had governance and oversight of the project. Participants gave recorded consent and were assured of anonymity and confidentiality. During interviews with matrons, it was evident that participants were experiencing a degree of distress so interviews with sisters and CNS/CRNs were conducted by researchers who had training in psychology and counselling so they could provide immediate support and signpost to the relevant expertise within the Trust to provide longer-term support.

### Data collection

Data were collected through semi-structured interviews conducted through online videoing software. The interview schedule with the operational leads focused on a description of the changes, what worked well, where there were challenges and what could have gone better, and what changes they felt should be implemented as business as usual. The interview schedule for the other interviews reflected on the changes in practice and how this had impacted on their work, how they led their teams (matrons and sisters), impact on the team and the personal impact. The interview schedule was reviewed by the Nursing and Midwifery Leadership Team. Interviews were recorded, notes were made from the interviews from the operational leads and the rest were transcribed verbatim.

### Analysis

The secondary analysis focused on aspects of the interviews related to leadership and were analysed using Framework analysis (Richie and Spencer 1994). Transcripts were coded for the overarching theme of leadership by two researchers and these sections were coded further to develop a framework. The framework was subdivided into the organisational levels of leadership so the interrelationships between decision and impact could be identified. A member of the evaluation team was a senior nurse within the Trust so a number of processes were implemented to limit bias. Four of the project team were not nurses but had extensive experience of working in healthcare and working alongside nurses so had insight into nursing practice. Analysis was undertaken by two people but the framework and subsequent charting were randomly checked by two others. Finally, an independent researcher with extensive qualitative health research experience reviewed the charting, subsequent interpretation and presentation of the findings. Members of the nursing leadership team identified the relevance to practice and implications for leading exceptional circumstances in the future.

## Results

A total of 48 members of staff participated in the evaluation (Table 1). The findings are presented as a summary of the Trust-wide operational changes followed by the themes that emerged from the interviews with the other three levels of leadership. For ease of reporting the terms Trust-wide, division, ward/department and individual are being used and more granular level detail is not provided to ensure anonymity.

**Table 1:** Summary of participants in the evaluation

| Group | Participants |
| --- | --- |
| Whole Trust (n=17) | Heads of the volunteer and patient experience service<br>Heads of staff experience/welfare<br>Leads for management of risk<br>Leads for Electronic Health Records and digital healthcare<br>Leads for nursing workforce<br>Deputy Chief Nurses |
| Division (n=7) | Matrons |
| Ward/department (n=8) | Sister/charge nurses<br>Senior CNS<br>Senior CRN |
| Individual (n=16) | CNS<br>CRN |

CNS: clinical nurse specialist; CRN: clinical research nurse

### Summary of the operational changes to the delivery of care

A summary of the changes made to the delivery of care across the Trust are presented in Table 2.

**Table 2:**
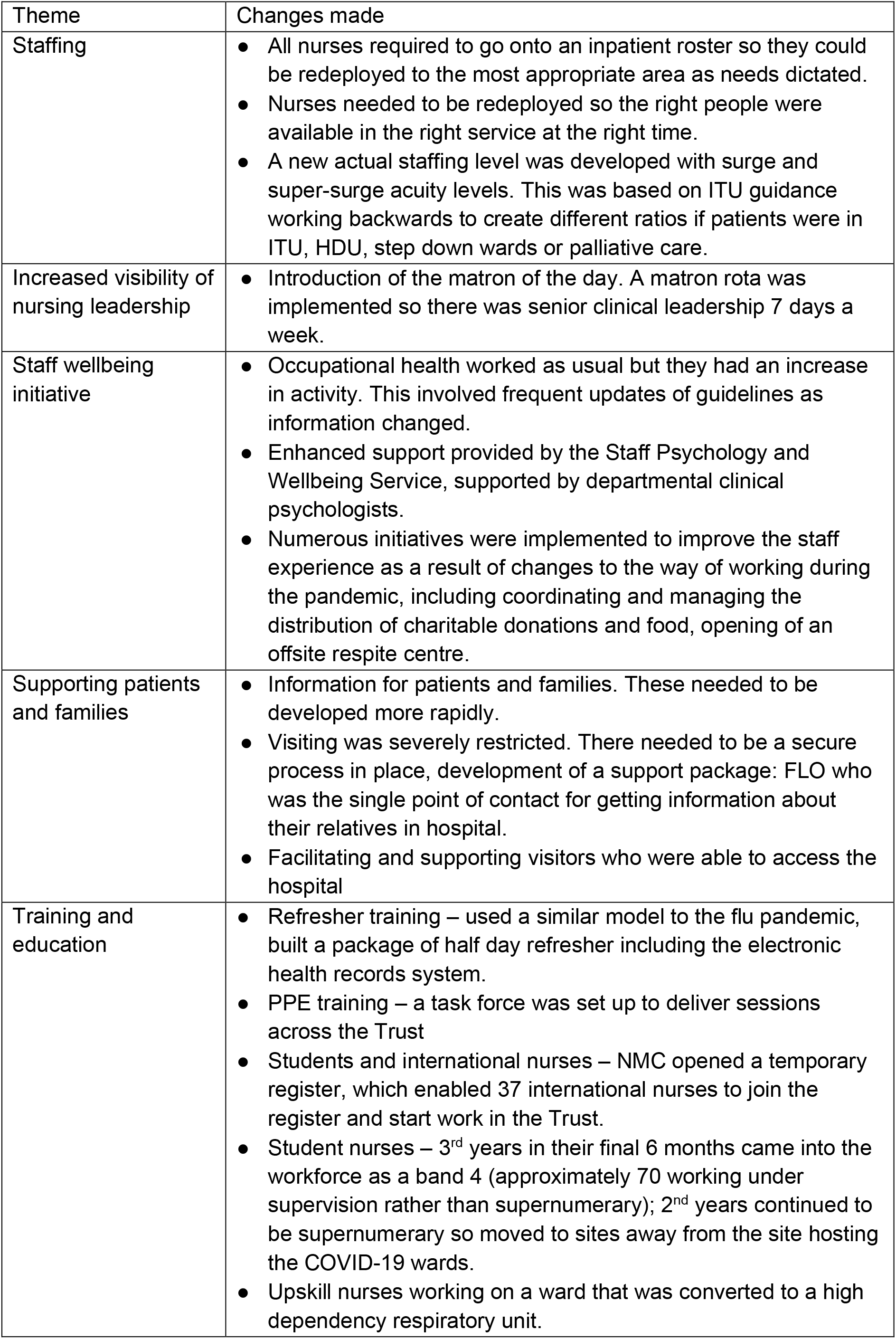
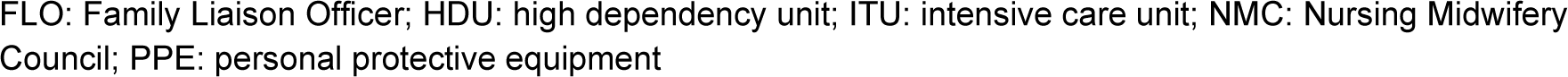
Changes made to accommodate the delivery of care during the pandemic FLO: Family Liaison Officer; HDU: high dependency unit; ITU: intensive care unit; NMC: Nursing Midwifery Council; PPE: personal protective equipment

| Theme | Changes made |
| --- | --- |
| Staffing | • All nurses required to go onto an inpatient roster so they could be redeployed to the most appropriate area as needs dictated.<br>• Nurses needed to be redeployed so the right people were available in the right service at the right time.<br>• A new actual staffing level was developed with surge and super-surge acuity levels. This was based on ITU guidance working backwards to create different ratios if patients were in ITU, HDU, step down wards or palliative care. |
| Increased visibility of nursing leadership | • Introduction of the matron of the day. A matron rota was implemented so there was senior clinical leadership 7 days a week. |
| Staff wellbeing initiative | • Occupational health worked as usual but they had an increase in activity. This involved frequent updates of guidelines as information changed.<br>• Enhanced support provided by the Staff Psychology and Wellbeing Service, supported by departmental clinical psychologists.<br>• Numerous initiatives were implemented to improve the staff experience as a result of changes to the way of working during the pandemic, including coordinating and managing the distribution of charitable donations and food, opening of an offsite respite centre. |
| Supporting patients and families | • Information for patients and families. These needed to be developed more rapidly.<br>• Visiting was severely restricted. There needed to be a secure process in place, development of a support package: FLO who was the single point of contact for getting information about their relatives in hospital.<br>• Facilitating and supporting visitors who were able to access the hospital |
| Training and education | • Refresher training – used a similar model to the flu pandemic, built a package of half day refresher including the electronic health records system.<br>• PPE training – a task force was set up to deliver sessions across the Trust<br>• Students and international nurses – NMC opened a temporary register, which enabled 37 international nurses to join the register and start work in the Trust.<br>• Student nurses – 3<sup>rd</sup> years in their final 6 months came into the workforce as a band 4 (approximately 70 working under supervision rather than supernumerary); 2<sup>nd</sup> years continued to be supernumerary so moved to sites away from the site hosting the COVID-19 wards.<br>• Upskill nurses working on a ward that was converted to a high dependency respiratory unit. |

There were two key themes that emerged from the data at division, ward/department and individual level: impact of leadership and impact on the delivery of nursing care.

### Impact of leadership

The leadership theme had five sub-themes: visible leadership; communication; accuracy and consistency of information; providing support; and impact of decision-making.

#### Visible leadership

There were varied views from matrons about the ‘matron of the day’. It was an important role because it extended cover across the trust to give an additional safety net. This increased visible leadership reducing bureaucracy and facilitated matrons spending more time in the clinical areas. However, some of the matrons felt that this role was unnecessarily and doubled up on work that was already being done. At ward/department level, visible leadership was not discussed, but individuals reported positively to having increased visibility of the matrons as they were more accessible, and individuals were able to communicate easily with them.

#### Communication

Matrons felt that matron-to-matron communication was better during the pandemic than it had ever been before. However, they felt that things were changing so fast that there was often no time to give explanations of changes to their teams. They recognised that this contributed to, and often fuelled their team’s anxieties and fears.

> *“I felt like a general in the first world war standing back sitting on a horse saying off you go, it’ll be okay. You can put your head up and climb out that trench and run across that no man’s land. It’ll be alright. And hands on heart not actually be 100% sure of what I’m saying, passing on is actually going to turn out kosher”(Matron)*

At ward/department-level, communication was sometimes reported as sub-optimal or inadequate, but sisters appreciated the circumstances they were in, and reaffirmed the view of matrons in understanding that the style of communication resulted from a state of crisis management, however, the process also created high levels of uncertainty. Late night communication from the operational team was felt to be done without concern for the impact this had on their team, either in being able to organise work or the emotional impact.

Conversely, individuals noted that the daily communications from the Trust were good and kept them informed of what was happening. However, the main changes that they did not feel was communicated well was around redeployment, especially from the managers who were leading it. There were inequalities in some of the consultations within their teams, with some being asked where they wanted to work while others were instructed that they were going to be moved without any consultation.

> *“There was no discussion. So whilst other members of my team were asked where they would prefer to work, and within the hospital, I was just told that this is what I was going to do. I had very little communication, my line manager did very little communication with me”* (Nurse)

#### Accuracy and consistency of information

Matrons felt as if there was an expectation that they were the expert, however they experienced rapidly changing information which made it difficult for them to be confident about the accuracy of the information they were distributing. An example of this was the inconsistency of the information around PPE, and the frequent change in recommendations. Information was often circulated without any forewarning, so matrons were not always prepared to be able to give the necessary support to their staff.

Sometimes they felt that the information was not always correct or accurate, the example cited was the notification that nurses were going to be redeployed immediately, but it was often days, weeks or months for this to be actioned. This waiting period prior to redeployment was a time of huge anxiety for their teams, which matrons subsequently needed to support. At ward/department-level, there was the same frustration as matrons about the constantly changing guidance, especially around PPE. While individuals reported good communication in the Trust, they initially felt that they were finding out information through social media, rather than their line managers. Nurses were given information at ward/department-level, but often got the impression that the people giving it knew as little as they did. Again, the inconsistent information about PPE was stressful, especially the constantly changing guidance.

> “*There was confusion about the masks, you know, whether we were the third of the mass or the FFP3 so in the end, someday, some nurses wore the surgical mask or some nurses wore the FFP 3 masks, you know, and someday some people wore the full the full surgical gown and other people didn’t you know, that that’s, that makes things a bit more stressful because it’s like, you know, people were doing what they felt more comfortable in. And that will actually maybe…we should know what we’re what we’re safe in*” (Nurse)

#### Providing support

Matrons saw it as their role to communicate with their team, so they were able to give them the appropriate support. Their increased visibility in the clinical area was important to facilitate this because it made them available, so staff could easily approach them. This was an important part of their role, and they needed to provide a lot of pastoral care to their teams, which was time consuming. This was also important for staff who were working from home, who equally needed their support, and the acknowledgement that their roles were as important as their colleagues, perhaps in part due to rhetoric at the time focusing primarily on staff working on the frontline.

Overall, at ward/department-level, redeployment of nurses was difficult. They did not always know where their staff were working or if they were off sick, so were not able to provide them with any support. There were pressures on some sisters to provide remote support to their team while they were not receiving any support themselves from the matrons. However, others reported feeling very well supported throughout. Individuals had a more negative perception of leadership support especially those who were required to deliver frontline care who had not delivered ward-based care for many years. They felt their fears were not taken seriously. Conversely, those who were working from home felt isolated by the focus on supporting nurses working on the frontline.

> *“The organisation, it’s all been about supporting people on the front line or supporting people coming in, and hasn’t been that much that has been geared to the people assigned to work at home …I feel a bit invisible”* (Nurse)

#### Impact of decision-making

Perceived poor decision-making at more senior levels impacted elsewhere. For example, matrons felt that the Trust-wide changes that were being implemented did not always align with the workforce requirements. They questioned whether there was enough nursing input into some of the decisions that were being made operationally. At ward/department-level, they questioned why the changes that were going to be required were not predicted at Trust-wide level and guidance implemented earlier. They felt that even when they were consulted about some changes, their input was ignored. At ward/department-level, concerns were expressed on several safety issues, especially redeployment of staff who had not done shifts for many years and the decision to move them into COVID-19 areas. The decision to move nurses was also often perceived to be very last minute, in a crisis management manner.

> *“The communication wasn’t great. There was rumours flying around left, right and centre before any of that actually happened. But I mean, I think that was a lot down to decisions not fully being made until, like, very last minute, I think our ward sisters were as honest with us as they could be”* (Nurse)

Finally, at ward/department-level, they felt that Trust-wide expectations were for all COVID-19 wards to function in the same way and did not take the existing ward culture into consideration, which sometimes resulted in conflict. Individuals understood that they needed to be redeployed but were confused and sometimes angry about how they had been notified. Some described an anxious and liminal period, having been given a short notice of redeployment followed by an extended period of time waiting to be deployed.

> *“The most stressful time when there was a letter sent… saying nurses will be redeployed, with immediate effects, but then nothing happens for another sort of eight or nine days after so, yeah, that was quite a tense period for everyone. For lots of my team, people I manage. Some were very fearful about where they’d go”* (Sister)

### Impact on the delivery of nursing care

The impact on the delivery of care was influenced by redeployment and teamwork.

#### Redeployment

The first challenge for the delivery of nursing care was allocation of shifts through the e-roster. The problem with the existing e-roster system was recognised Trust-wide; however, the fact that a new e-roster was not in place at the time of the pandemic compounded the challenge of redeployment. At ward/department-level, it was not until redeployed nurses were allocated a ward that they would know the shifts that were being covered. While individuals were able to organise their shifts, they were not organised in advance in the same way as non-deployed nurses, which had an impact on their personal life. The biggest challenge was ensuring there were enough nurses available to deliver care. The problem with the e-roster resulted in matrons not always knowing who was on duty.

They had organised redeployment early in the planning stages based on their relationships and knowledge of their staff – they knew their teams’ personal histories so matched decisions to best fit their emotional situation. This process was therefore undermined by the limitations inherent to the e-roster, which may account for some of the issues reported by staff during this period.

The matrons reflected that the general epistemic uncertainty surrounding COVID-19 as the situation developed resulted in the Trust being over prepared to a degree: *‘Prepared for a war that didn’t come*’. Consequently, there were lots of staff who were redeployed, who did not necessarily need to be redeployed. One of the issues noted by matrons was a lack of clarity Trust-wide on the process for transitioning redeployed nurses back into their own roles. This was a frequent enquiry at ward/department-level and from individuals, who felt they were unable to answer this. Sisters felt that when it was clear that staff were not needed, nurses should have been able to go back to their own roles.

> “*There was a big call to people to be deployed, but we had to ask for them to be able to come back and I think, and, you know, we were getting calls from people saying there was no work and that they had several nurses per patient and on some areas, and I think it would have been helpful for that to be made more explicit*” (Sister)

Some of those at ward/department-level found that their whole team were redeployed resulting in leading a new team of redeployed nurses. A particular challenge noted was redeployment resulted in the workload of redeployed nurses being distributed to those who were not redeployed because care for existing patients still needed to continue. At ward/department-level they needed to provide a lot of support for redeployed nurses, especially those who had not been ward based for a long time experienced a lot of anxiety. Similar to matrons, they noted the transition back was often more problematic. They found that they were given no notice or details about when their nurses were transitioning back.

> *“Think maybe just for warning the nurses on the wards that it looked like they weren’t going to be needed much longer. And they have to think about the fact they may soon be going back to their original role was what happened. They turned up to the ward one day and then we’re told we don’t need you anymore. And that was that” (Sister)*

Individuals had a lack of choice on who worked from home, and who was redeployed. Some got the impression that BAME staff were less likely to work at home and more likely to be redeployed to frontline COVID-19 care. Individuals who were given a choice viewed this situation more positively than those who were not. The Trust being over prepared, noted by matrons, was an observation reinforced by individuals who had been redeployed, who found that often there were more staff on duty than patients. They questioned why they could not return to their roles so they could continue their work. Some of the nurses who were redeployed reported that they were not always treated very well and felt that the decision on where to deploy them was made based solely on their clinical background. This reflected the reports from matrons on the importance of knowing the personal history; for example, having to go back into critical care had a negative impact on nurse’s emotional wellbeing.

> *“They didn’t really think about you as a person when they decided to place you in ICU, they just saw your background” (Nurse)*

Though initially there was a willingness to be redeployed, many felt that after their experiences of being redeployed, they may not be as willing to be redeployed again during a second wave.

> *“I’m telling you honestly, I cannot do this again. I’ve done it once. I would, I wouldn’t be able to do it again. I had to do it. I did it. But if I have to do it again, my God, it would be very, very challenging” (Nurse)*

Again, the transition back to normal roles was another period of anxiety and uncertainty and nurses found that when they did transition back, it was not transitioning back to normal, so they still felt as if they were in *‘limbo’*. Often after they had transitioned back, they found that no one had supported their work while deployed, leaving them with a lot more work to catch up following the turbulence of their redeployment. Nurses who were working from home had additional pressures due to the perception that they should be more freely available while not on the frontline:

> *“When I’m working from home, it’s like there’s more pressure; you should answer your email straightaway I, should deal with this right away… They sort of expect you to do more things and to do it very quickly. And to be always, you know, on top, so I can’t leave my computer because at nine o’clock somebody emailed me emailing me already and they expect an answer” (Nurse)*

#### Teamwork

Firstly, the matrons felt that they worked very closely together, which provided them with a lot of support. At ward/department-level, participants also reported that having their team together was important for support; it was felt that it was better to care for a different population than splitting the team. Keeping the team together was enhanced because of the availability of virtual communication, so nurses working from home were able to benefit from the team support. However, ward/departments who were given a new team to manage, led to feelings of isolation. A new team resulted in a change in the dynamic; when non-nursing members of the team tried to enforce a culture from their previous location, this caused conflict. Where teams had been split up, at ward/department-level the changed dynamic needed to be managed when the team returned because there was resentment from nurses who were COVID-19 facing when they met members of their team who were not, who were viewed as having worked in a ‘*bubble’*. However, a positive aspect of the change in the ward team was the lack of role definition so it was a lot easier to manage patient care because everybody was willing to do everything.

> *“The ward teams were so lovely and everybody just banded together and I’m just so proud to be part of that”* (Sister)

Nurses felt that the mechanisms that they and their sisters had put into place to develop a team spirit worked well, for example team WhatsApp groups. However, some of the nurses who were redeployed reported having no line manager contact throughout their redeployment, so they felt very isolated.

> *“I just felt that the way I was treated my line manager, not one day from when I was redeployed, ever made contact with me to say, how are you doing? You know, is everything okay? Not once, never heard from her” (Nurse)*

There was a general sense that there was a flattened hierarchy and a sense of community, ‘*everybody was in the same boat’*; medical colleagues were contributing to the delivery of fundamental nursing care that COVID-19 patients required. This flattened hierarchy was helpful for facilitating support for redeployed nurses; grades became irrelevant when seeking and giving advice and guidance. Nurses working from home especially missed being with their team and missed their administration staff, who would provide them with a lot of support pre-lockdown. However, nurses were not always welcomed into the teams they had been redeployed to, with examples given of host nurses acting as if they were superior and not acknowledging that those who were redeployed were often experienced and highly qualified nurses.

> *“I felt like the team was not ready to accept us…I understand about that culture…it’s a culture inside [CLINICAL PLACE] that […] nurse just felt you’re better than the other nurses*.*”* (Nurse)

## Discussion

This study reports the impact of leadership on the nursing workforce of the transformation of the workplace to accommodate the COVID-19 pandemic. Previous studies have highlighted protective measures that organisations should implement to minimise the psychological burden on clinical staff of responding to a pandemic. These included clear communication, leadership and access to PPE, as well as access to psychological support, provisions of food and necessities, adequate shift patterns, options of alternative accommodation and access to up-to-date training and education (Kisely et al., 2020). In this single centre study these recommendations were taken up as policy decisions that were enacted by nurse leaders. The initial evaluation, commissioned by the organisation, was designed to describe the changes that were made with the intent to identify which were beneficial and could be used in future waves or crisis situations and which should be implemented as business as usual. This further analysis has highlighted some key themes about how nurses responded to these changes.

### Communication

Clear communication has been identified as a protective factor (Kisely et al., 2020) and in this study nurse leader participants described a rapidly changing scenario where they were often unsure of the accuracy of the information they were sharing. This was occurring in the context of a rapidly evolving international pandemic and a national response where information was emerging and changing equally rapidly. Perhaps unsurprisingly consistency of information was identified as a theme in this analysis where the fear that they were not communicating accurate information was a source of concern for nurse leaders. Participants reported a wide variety of experiences in the communication about decisions that affected them as individuals. While the Trust-level information published regularly was reported positively, communication interactions about what to do or not to do was reported as inconsistent with participants having both positive and negative experiences.

For many there was an expectation that those ‘higher up’ should know more and tell them clearly what to do. This expectation resonates with the need for clear communication as a protective measure reported in other studies (Kisley et al., 2020), but also highlights the challenges of providing this clear communication in a rapidly evolving situation. The ideal of policy decisions that are taken up and communicated in the same way across a large organisation by a multiplicity of individuals, in multiple different situations, has been challenged by several organisational theorists (Stacey 2007, Shaw 1997, Mowles 2011).

However, this expectation remains central to systems thinking which dominates in healthcare (Kaplan & Norton, 1996, Drucker 1996, Argyris & Schon, 1978). In this study participants brought this ideal as an expectation to their work in the pandemic and their experience did not match the ideal. Arguably this added to the moral distress experienced as this ideal was not the reality of what was happening.

The theme of communication demonstrates clearly how participants at all levels experienced the uncertainty they were working in as difficult. Their experience of changes during the pandemic were not the direct translation of policy decisions in a linear way top to bottom as systemic thinking leads them to believe it should be but instead the result of a multiplicity of human interactions.

### Decision-making

Findings show that there was variation in how policy decisions were taken forward and in local decisions that resulted from them. This was particularly evident in the work of redeployment; previous studies have illustrated the negative impact of uncertainty about deployment in a crisis (Li et al., 2017, Dunn et al., 2020) and the current study illustrates the challenges of redeploying staff in an unfolding crisis where matching demand and need is unpredictable. A policy decision to redeploy staff was conveyed in a letter to all nurses and midwives and this was translated into processes that included the implementation of a matron roster, development of a new actual staffing level (ASL) tool and the creation of new teams on the electronic rostering system. However, findings show that the way staff were redeployed was not achieved through the translation of policy into practice in a linear way but in fact redeployment was a messier exercise. Staff at all levels described their individual experience of this policy decision in both positive and negative terms and the evidence shows that the policy and processes were adopted in numerous different ways in different circumstances.

Matrons reported that their redeployment decisions were based on their relationships and knowledge of their staff – they knew their teams back stories so made decisions to best fit their individual situation. Thus, the decision about redeploying staff was reached not through the application of rules laid out as policy in the redeployment letter or through the new processes created in the ASL or the e-roster, but through the exercising of practical judgement by nurse leaders in numerous different situations. Nurse leaders reported difficulty in bringing the organisational expectation laid down in policy together with the expectation of nurses and their own values of caring for staff and patients to make decisions. This balancing of the competing demands of the organisational policy with the needs of staff has been reported previously as a difficult but core element of daily nurse leadership (Phillips & Norman 2020). Arguably the work and the impact of these decisions was greater in this case where the language used illustrates the moral distress experienced when making and communicating these decisions:

> *“I felt like a general in the first world war standing back sitting on a horse saying off you go, it’ll be okay. You can put your head up and climb out that trench and run across that no man’s land. It’ll be alright. And hands on heart not actually be 100% sure of what I’m saying…” (Matron)*

While the technical limitations of the digital systems for deploying staff were widely reported, analysis also show the limitations of a digital system alone for redeploying staff. This is because they demonstrate that leadership decisions by matrons and ward sisters were not a direct translation of organisational policy or tools like a roster system but the result of bringing knowledge about and the expectations of affected individuals into view alongside the need for organisational need for safe staffing. This practical decision-making is the professional judgement of the experienced nurse leader and the complexity and impact of such judgements have the potential to lead to the moral distress that has been reported in previous studies (Phillips & Norman 2020, Ulrich 2014)

The findings illustrate the impact of policy decisions on all participants; what was intended as a beneficial change when setting policy was experienced as both positive and negative across the organisation and this was situation dependent. This presents a challenge to existing thinking about command and control approaches to crisis management and may account for why some of the recommended protective measures did not always seem to afford protection to individual staff wellbeing in this case.

### Nurse leadership

It has been reported that a lack of clear leadership in times of crisis can lead to an increase in psychological distress for nurses (Li et al., 2017, Mao et al., 2018). In this case the need for leadership from those above was widely reported and uncertainty was reported by nurses in all roles at all levels as they sought to be prepared but had to respond to an unpredictable unfolding crisis. They reported a need for predictability, early planning and early decisions and described looking to leaders in the hierarchical structure for direction while recognising who did not have answers and were in the same situation.

Findings demonstrate the challenges of providing consistently clear leadership for matrons who reported that information was moving so fast that they were unclear of the accuracy of what they were conveying. They believed there was an expectation of them to provide clarity from junior nurses who conversely reported that they did not expect this because they recognised the rapidly changing nature of the situation. Regardless, the belief that this expectation existed was a source of concern for matron leaders. Despite this concern and the challenges experienced by all participants the visible clinical leadership of matrons was widely reported as positive by frontline staff. Matrons themselves reported that they felt supporting nurses was a key element of their role and while they themselves may have felt ill equipped findings show this was a beneficial element of the organisational response

### Teams

The pandemic response required the establishment of new teams to staff new areas and increasing the size of teams for areas most affected like intensive care. Responses demonstrate the value that is placed upon team as a means of support and this is in keeping with previous studies that show peer support as a key protective measure for those working in crisis situations. However, experiences of team-working were reported with wide variation. Matrons reported working much more as a team and described this as positive while for others they had varied experience of being welcomed or not into new teams. For some no longer being part of a team was a challenge and led to feelings of isolation.

This poses the question if peer support is a protective measure for staff wellbeing in a crisis situation how this can be best achieved when the crisis itself requires the breakdown of existing teams and establishment of new teams. The work of healthcare organisations has been described as a negotiated order (Mauksh 1973) and this negotiation points to our dependency on each other. Many participants talked about culture when referencing teams. How is culture rapidly established in a new team? How does it change and how are cultural practices negotiated over time? The cultural shift as a result of the pandemic warrants further exploration.

The current study had a number of limitations. Firstly, this was secondary analysis of existing data. While leadership emerged as a key factor, the interviews were not specifically about leadership and therefore more in-depth probing was not undertaken. More detailed understanding of the challenges in leadership could therefore be missed. Second, individual nurses included CNS/CRNs, who were band 6 or 7 and although they were mostly redeployed, they were all experienced nurses. The study does not account for the experiences of nurses who continued on their wards, those who were band 5 or below, and international nurses and students on the temporary register. Finally, there were no matrons, sisters or nurses from critical care, only those who were redeployed to these areas so only one perspective is presented. However, despite these limitations this is the first study reporting on nursing leadership during the pandemic. While there are numerous publications focusing on the importance of leadership, these are mostly editorials, commentaries and opinion pieces (Acquilla et al. 2020, Hoffman et al. 2020, Rosser et al. 2020, William et al. 2020); our analysis has provided evidence to support the importance of nursing leadership and the impact this can have on the nursing workforce. Data reflected experiences across numerous roles and grades in the nursing workforce and has provided an in-depth view of nurses lived experiences of leadership during the pandemic. Furthermore, data were collected during and shortly after the peak of the first wave of the pandemic in 2020 so there was less recall bias.

### Recommendations

The findings of this study support the claims in previous studies that there are protective measures that should be implemented to support the wellbeing of nurses working in crisis situations. However, they also point to the challenges in implementing these measures.

Arguably these challenges cannot be eliminated given the inevitable fact that they emerge from the practice of implementing change among interdependent individuals in healthcare crises. Therefore, a key recommendation of this study is that wellbeing initiatives where staff have the opportunity to reflect on the work they do together are implemented. Such measures have the potential to enable nurses to make sense of their experience and explore any moral distress that results from working with the increased ambiguity and uncertainty that is inevitable in crisis situations. Furthermore, while the experience of communication was reported as variable, for the reasons explored, two elements stood out as consistently well reported: visible clinical leadership of matrons and organisation wide communications.

These tools have the potential to provide the clear communication and leadership needed in a pandemic.

## Conclusion

The NHS has never experienced a situation of this magnitude before, so leaders did not know what to expect. Reports from China and across Europe suggested there would be a deluge of sick patients and therefore significant changes were made to the hospital environment and workforce to accommodate this. These changes reflect the protective changes recommended in previous studies of crisis situations in healthcare (Barello, Palamenghi, & Graffigna, 2020, Ma et al., 2018, Maunder at al., 2008, McAlonan et al., 2007, Nasrabadi et al., 2003). This study has highlighted how these changes were taken up in different ways by nurses and nurse leaders in different situations who experienced them in both positive and negative ways. It is important to recognise that the challenges and distress nurses experience when making moral decisions about what to do and what not to do in the complex world of healthcare is not new. Consequently, ongoing support for nurses’ well being will remain critically important beyond the pandemic. In the initial evaluation the research team were asked to identify what should be taken forward as business as usual; findings demonstrate the need to support nurses in making sense of their situation together. It has been argued that this is best achieved through creating reflexive spaces where they can think and learn together with supporting the development of skills and resilience for working in complexity (Phillips & Norman 2020, Maben 2020).

### Epilogue

The ‘*war that did not come’* arrived in December 2020 and the importance the Trust placed on learning from the first wave of the pandemic facilitated a different leadership approach. At the heart was communication, choice and collaboration. There was a need for large numbers of staff to move to the critical care units and surge areas to support the unprecedented numbers of patients requiring intensive care or enhanced respiratory support and assist to rapidly roll out the vaccination programme across [location to be added after review]. The observation in the first wave that many staff were willing to be redeployed supported a volunteer programme and therefore staff had the choice to work in critical care/enhanced care or with the administration of vaccines. This enabled some control over the shifts they worked and were able to balance redeployed work with their existing caseloads so their patients continued to receive support. The hospitals across [location after review] collaborated so there was sharing of resources and expertise, and movement of patients to ensure workloads were distributed evenly across the patch. Finally, the daily communication implemented during the first wave was expanded to include a weekly all staff virtual briefing where the Chief Executive, Chief Nurse and other members of the senior leadership team could update staff on what was happening across the Trust and answer questions. This was regularly attended by over 700 members of staff (plus those who accessed the recording on the Intranet). This was particularly important for the large numbers of staff required to work at home so they remained connected to the Trust.

While this epilogue is from the perspective of nurse leaders and researchers who observed the process, it may not reflect the experiences of those nurses whose work was impacted by leadership decisions. However, learning from wave 1 we are confident that our recommendations and conclusion apply now: it is essential the wellbeing of our nursing workforce is prioritised to ensure they have the emotional resilience to be able to deliver care to the anticipated high numbers of patients who are going to require treatment as we move forward.

## Data Availability

No data are available

## Acknowledgements

We would like to thank the Nursing and Midwifery Leadership Team for commissioning and supporting the evaluation these data were derived from. We would also like to thank all the participants for giving us their time during the pandemic and for sharing so honestly about the impact of working in a pandemic.

